# Design and implementation of a multi-country targeted health facility assessment to measure integrated, people-centered, quality of services

**DOI:** 10.64898/2025.12.16.25342344

**Authors:** Shivam Gupta, Japneet Kaur, Rohit Jain, Alexander K. Rowe, Pankhuri Jha, Rasheed Raji, Arjun Arakal, Chris White, Marasi Mwencha, Shunsuke Mabuchi

**Author notes:** Corresponding Author Dr. Shivam Gupta, Associate Professor, Department of International Health, Johns Hopkins Bloomberg School of Public Health, Baltimore, MD 21205, USA.

## Abstract

**Introduction:** Integrated, people-centred, high-quality services are essential to achieving universal health coverage. Traditional health facility assessments, while valuable, often focus on structural inputs and are resource-intensive, limiting their frequency and programmatic utility. The Global Fund to Fight AIDS, Tuberculosis (TB), and Malaria, developed the Targeted Health Facility Assessment (tHFA) to inform country plans and monitor progress on the Global Fund Strategy (2023–2028) of maximizing people-centred integrated systems for health. This paper describes the development, methodology, and implementation of the tHFA baseline round and discusses key issues related to institutionalizing these assessments to monitor service delivery process and outcomes to improve health services in low- and middle-income countries (LMICs).

**Methods:** The tHFA is a cross-sectional, multi-country facility assessment developed to support countries and monitor progress of four key performance indicators in the Global Fund’s strategic monitoring framework and seven programmatic indicators for the Global Fund’s investments to strengthen horizontal programs. In 2024, the Global Fund implemented a baseline survey across 2,272 facilities in 18 LMICs using a modular questionnaire covering integrated delivery of antenatal care, HIV, TB and malaria services, supportive supervision, community health worker (CHW) support, and patient-reported experience measures. Probability sampling was used to select 120 health facilities per country, and data were collected through direct observation, health worker interviews, and record reviews. Post-stratification weighting accounted for facility replacement and other sampling deviations. The Global Fund embedded data quality assurance through training, pilot testing, and real-time monitoring.

**Conclusion:** The tHFA provides a pragmatic, agile and scalable approach to monitor integrated service delivery and people-centred quality of care in LMICs. Its focused scope on service delivery processes and outcomes, inclusion of CHWs, and alignment with the Global Fund’s strategic objectives make it a valuable tool to guide decision-making, inform grant monitoring, and support health system strengthening efforts across LMICs.

## Introduction

Despite substantial progress in health service accessibility across low- and middle-income countries (LMICs) since 2000, actual health service delivery remains fragmented with quality deficits, resulting in low health system responsiveness and poor user satisfaction.[1,2] In 2015, all United Nations Member States adopted the 2030 Agenda for Sustainable Development, which contains 17 Sustainable Development Goals (SDGs). The broad scope of SDG 3 (“Good Health and Well-Being”)—particularly, Target 3.8 (“Achieve universal health coverage [UHC] by 2030”), requires global health policymakers, donors and program implementers to go beyond service access and improve what happens once people encounter health services.[3,4] This call to improve the process of service delivery was echoed in the World Health Organization’s (WHO’s) World Health Report 2013 and in the subsequent WHO Framework on Integrated, People-Centred Health Services (IPCHS) in May 2016, emphasizing integrated, high-quality, patient-centred services at all levels of care.[5,6]

The IPCHS framework emphasizes the transformation of health services to ensure they are accessible, comprehensive, coordinated, and responsive to people’s needs and preferences. The Lancet Global Health Commission on High Quality Health Systems further highlighted that improving health system performance requires health facility assessments (HFAs) that are agile and focus on the process and outcome of care rather than inputs.[2]Concurrently, the Primary Health Care Performance Initiative (PHCPI) contributed significantly to the understanding that achieving quality, sustainable UHC necessitates a shift from vertical programming towards integrated health systems, with integrated service delivery as a key component. The WHO Operational Framework for Primary Health Care formalized this perspective.[7]

Since its inception, the Global Fund to Fight AIDS, Tuberculosis (TB) and Malaria (Global Fund) has invested in strengthening the horizontal investments in health systems as an enabler for its disease-focused programming. Realizing the unique opportunity for action presented by the SDGs, and national commitments to UHC, and building on renewed attention on primary health care (PHC) catalysed by the 40th anniversary of Alma-Ata Declaration of 1978 and the Declaration of Astana in 2018 the Global Fund Strategy (2023‒2028) elevated the achievement of “maximizing people-centred, integrated systems for health to deliver impact, resilience, and sustainability” as a key objective across all HIV, TB, and malaria (HTM) investments.[8,9]This strategic shift aims to build on the health systems strengthening investments made in response to the COVID-19 pandemic while contributing to the SDG target of UHC through strengthened health systems. Monitoring Global Fund investments to strengthen health systems via integrated, people-centred quality services (IPCQS) has been operationalized via a framework that includes key performance indicators (KPIs) to measure integrated service delivery, service quality including patient-reported experience measures (PREMs), and support to community health workers (CHWs).[10]

Monitoring the Global Fund’s progress toward IPCQS over the five-year strategy period requires regular data collection on service delivery processes and outcomes at the point of care, using a baseline of 2024-25 to coincide with the signing of new Global Fund country grants, followed by regular rounds of HFAs. A review of available HFA questionnaires, including the Service Availability and Readiness Assessment (SARA), Health Facility Census (HFC), Service Provision Assessment (SPA), and the WHO Harmonized Health Facility Assessment (HHFA) revealed an emphasis on measuring structural readiness and a lack of data elements on people-centred outcomes and support to CHWs.[11] While SPA and HHFA include modules on process of care and patient satisfaction, historically these assessments have required extensive planning and are never conducted in multiple countries simultaneously at regular intervals. Using the available questionnaires as a starting point, the Global Fund designed a Targeted Health Facility Assessment (tHFA) and implemented a baseline round in 18 LMICs in 2024-25. This paper describes the development, methodology and the implementation of the baseline round of the tHFA and discusses key issues related to institutionalizing agile and focused facility assessments to monitor service delivery processes and outcomes to improve accountability for service integration and quality of health services in LMICs.

## Methodology

The baseline round of the tHFA is a cross-sectional, multi-country health facility survey designed to address the persistent gaps in monitoring of health systems strengthening by focusing on the process and outcome of service delivery. Its main objective is to measure 11 quantitative indicators that reflect the Global Fund’s strategic priority of integrating delivery of vertical (e.g., HTM) and horizontal (e.g., workforce, laboratory equipment, supervision) programs at the point of service delivery for an integrated, high-quality experience for the patient.

### Questionnaire Development

The Global Fund developed the tHFA questionnaire by systematically mapping existing questions from the WHO HHFA, filling gaps through a targeted literature review, and consulting with technical experts who participated in developing the strategic monitoring framework for the Global Fund. The group of experts helped with the technical validation and enhanced the specificity of these indicators, aligning them with the frameworks from the WHO and the World Bank that emphasize service delivery integration and quality of care. The consultation also included several Global Fund Secretariat teams including country teams for the 18 countries, three disease (HTM) teams, cross-cutting thematic teams (laboratory, human resources, community rights and gender, supply chain, and pandemic preparedness and response), the ministries of health of these countries, and their national disease programs. A distinctive feature of the tHFA is the inclusion of PREMs and support for CHWs within the assessment questionnaire, both of which are Global Fund strategic priorities.

The baseline round was carried out in 2024-25 across 18 countries: Burkina Faso, Cameroon, Chad, Congo, Democratic Republic of the Congo (DRC), Gambia, Ghana, Guinea-Bissau, Liberia, Madagascar, Malawi, Mali, Mozambique, Niger, Nigeria, Tanzania, Senegal and Uganda. The tHFA measures 11 quantitative composite indicators, including four KPIs (Table 1: Section 1) and seven programmatic indicators (Table 1: Section 2).[10,12] The seven programmatic indicators monitor progress on the Global Fund’s investment priorities at the facility level to strengthen horizontal programs of human resources, laboratory diagnostics, functional oxygen availability, community-led monitoring, and availability of disease guidelines and treatment completion rates.

**Table 1:**
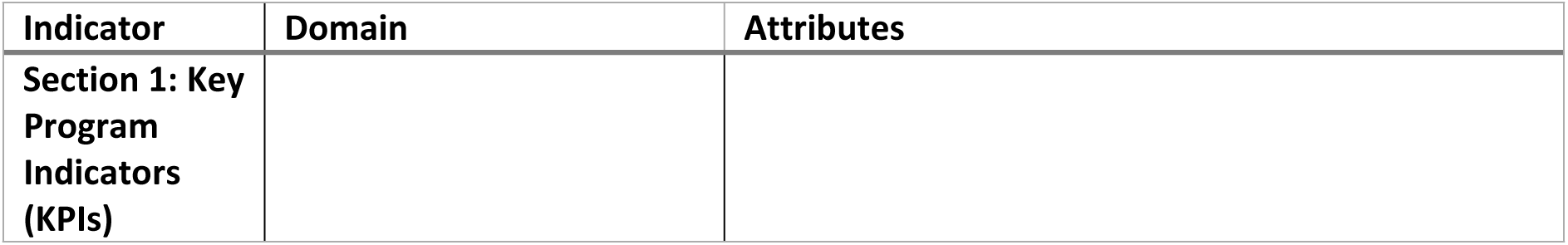
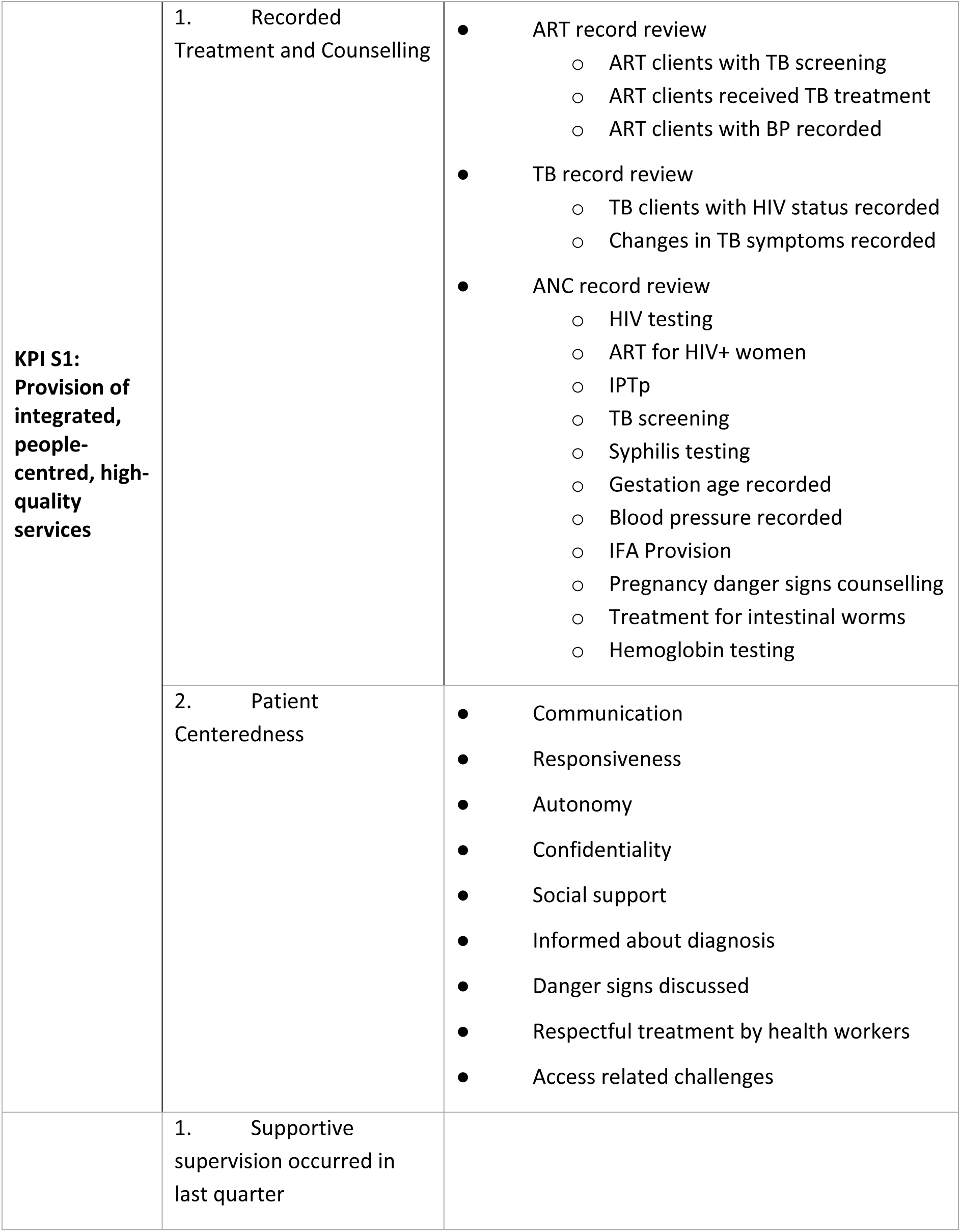
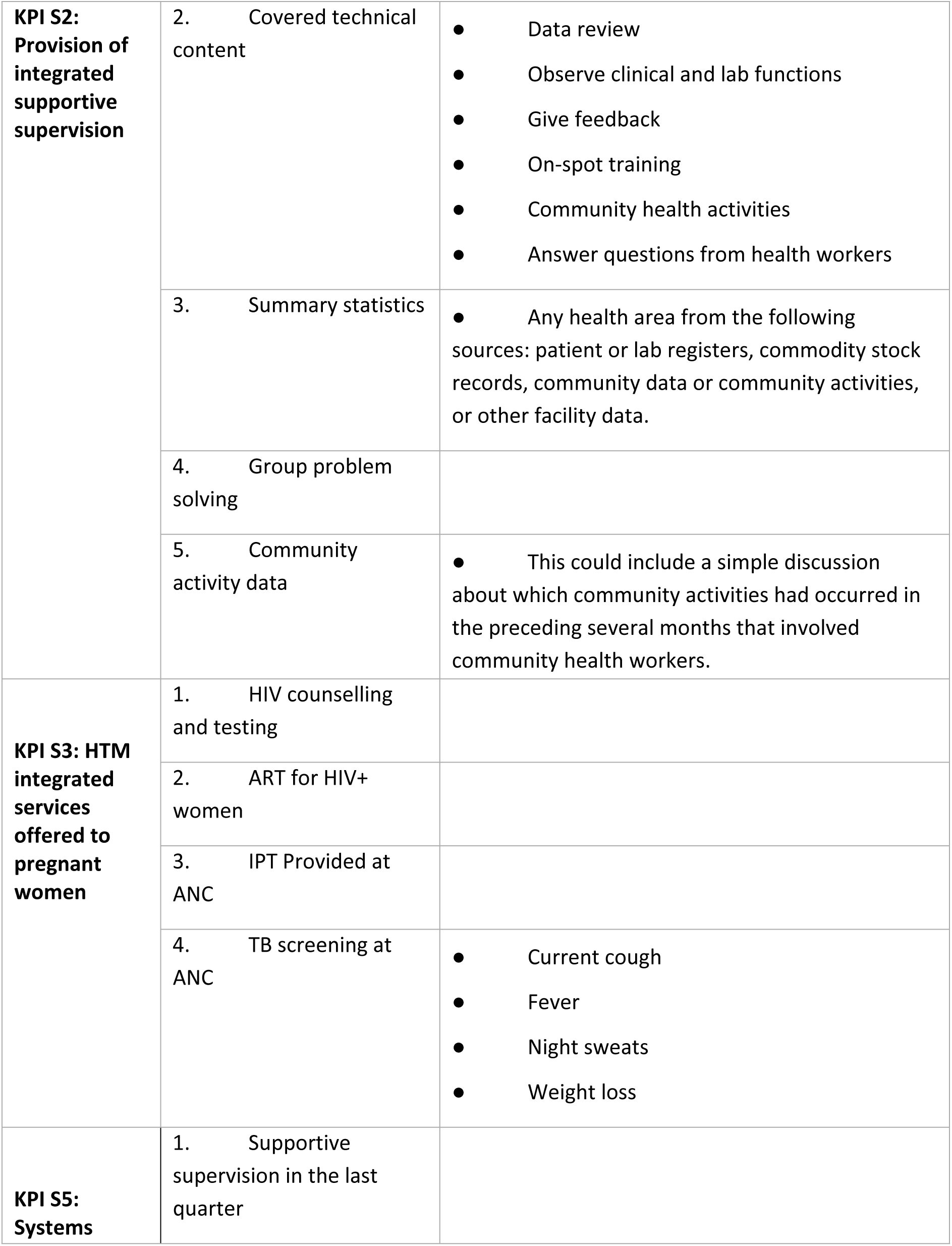
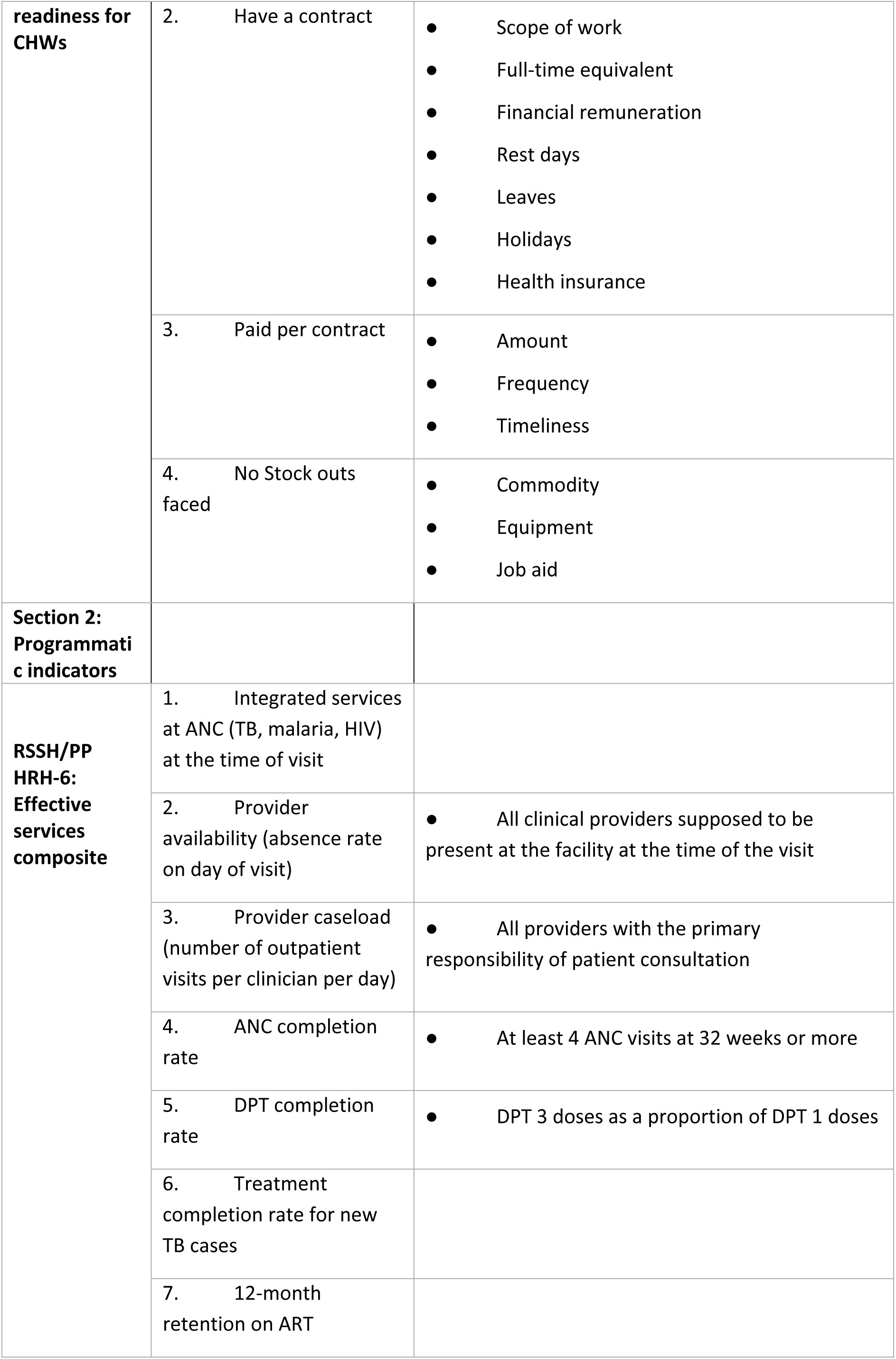
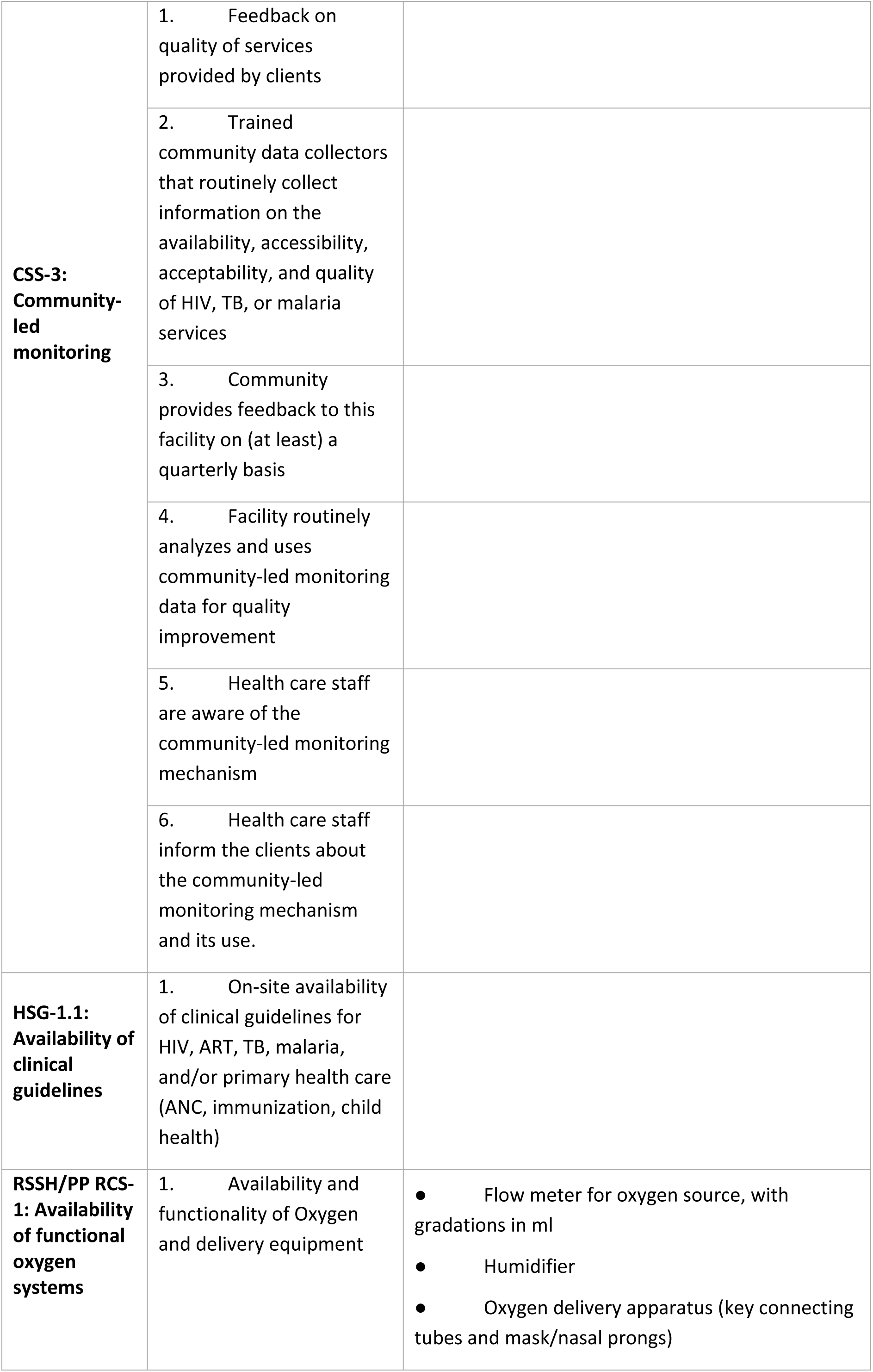
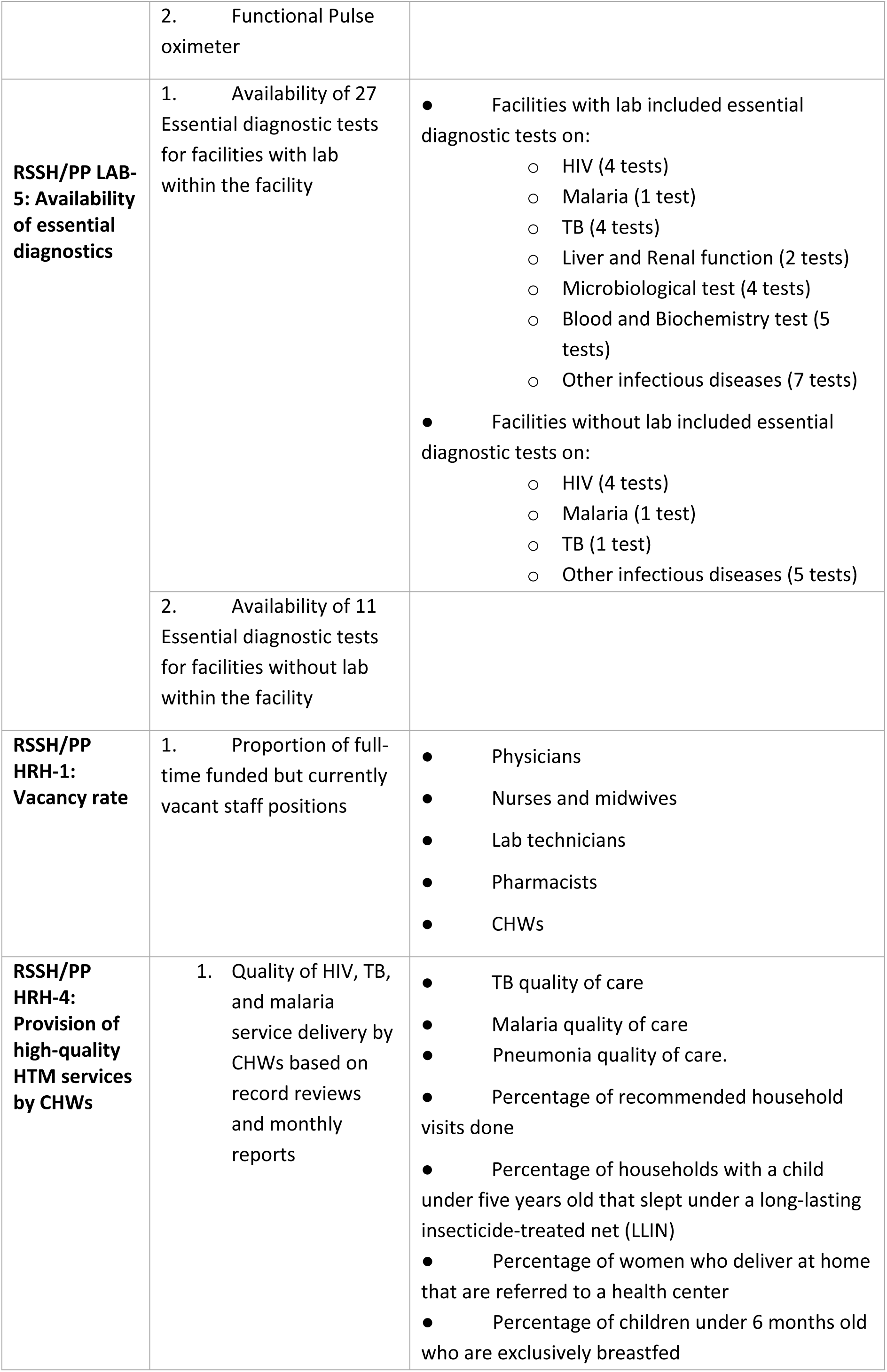
Domains and Attributes of the 11 Indicators measured through Targeted Health Facility Assessment (tHFA)

The 11 indicators align with the Global Fund’s strategic monitoring framework and follow the terminology and coding system that corresponds to the thematic investment areas of the Global Fund Strategy 2023–2028. Each indicator is designed to capture service delivery processes (integrated service delivery and quality of care) and outcomes (patient-reported experience), providing a pragmatic way to track health system performance across multiple countries.

#### 1. KPI S1: Provision of Integrated, People-Centred, High-Quality Services

This indicator measures the delivery of high-quality, integrated services at health facilities, incorporating technical and interpersonal aspects of care. It moves beyond input-based metrics to capture what happens at the point of care, emphasizing adherence to clinical protocols and respectful communication with patients. This indicator has been developed with two key dimensions: (i) health worker (HW) competence/treatment and counselling provided to patients for sexual and reproductive health (SRH), antenatal care (ANC), and HTM delivered in an integrated fashion; and (ii) patient centeredness-that is, patient-reported experience including communication, respect, autonomy, confidentiality, and social support. Data for the first dimension are collected via review of patient records including ANC, ART and TB records, and the data for second dimension are collected through patient exit interviews.

#### 2. KPI S2: Provision of Integrated Supportive Supervision

This indicator measures the components and quality of supportive supervision provided to HWs at the facility level. The rationale recognizes that routine, structured supervision contributes to higher adherence to clinical guidelines[13] improved service integration, and stronger staff motivation. In many settings, however, supervision remains irregular, disease-focused in scope, or punitive rather than constructive. KPI S2 captures the frequency and scope of supervisory visits, including whether they cover multiple service areas (HTM), review records, offer feedback, and support group problem-solving. Data for this indicator are collected through interviews with the facility in-charge.

#### 3. KPI S3: HTM Integrated Services Offered to Pregnant Women

This indicator measures whether pregnant women receive a coordinated package of essential services across HTM domains during ANC visits. The rationale grounds the understanding that pregnancy presents a critical opportunity to deliver multiple life-saving interventions, and fragmented service delivery can lead to missed diagnoses and preventable adverse outcomes. Components include whether the facility provides HIV counselling and antiretroviral therapy (ART) for HIV-positive patients, intermittent preventive treatment of malaria in pregnancy (IPTp), and TB screening as part of routine ANC. Data for this indicator are collected through review of ANC records.

#### 4. KPI S5: Systems Readiness for CHWs

This indicator assesses whether health systems are structurally equipped to support CHWs who play a vital role in delivering primary care at the community level. The rationale recognizes that well-supported CHWs improve coverage and quality of services, especially for marginalized populations. However, many systems lack formal mechanisms to adequately train, compensate, and supervise them. KPI S5 measures the availability of formal contracts with detailed terms of reference, supervision mechanisms, availability of supplies including key equipment, job-aids and commodities, and timely remuneration. Data are collected through structured in-person interviews with CHWs who are invited in advance to sampled facilities.

#### 5. Programmatic Indicator 1: Resilient and Sustainable Systems for Health (RSSH)/Pandemic Preparedness (PP) Human Resources for Health (HRH)-6: Facilities Providing Effective Services

This indicator measures a facility’s service delivery capacity by combining performance across seven sub-indicators. These include: (i) provider/staff availability on the day of visit, (ii) provider case load (number of outpatient visits per clinician per day), (iii) ANC completion rate, (iv) TB treatment completion rate, (v) Diphtheria, Pertussis, and Tetanus (DPT) vaccine completion rate, (vi) HTM integration at ANC, and (vii) 12-month ART retention rate. The rationale provides a snapshot of how effectively a health facility functions in delivering basic primary health services Data are collected through record reviews and HW interviews.

#### 6. Programmatic Indicator 2: Community System Strengthening (CSS)-3: Community-Led Monitoring

This indicator gauges the presence of community-led monitoring mechanisms within the health facilities delivering services. The rationale stems from the Global Fund’s investment strategy to promote accountability, community ownership, and data-driven advocacy, which can improve service responsiveness and equity. Components include confirming the existence of structured community-led monitoring processes (e.g., service user feedback mechanism), independent community leadership of these efforts, and integration of findings into facility improvement plans. The data are collected through an interview of the facility in-charge.

#### 7. Programmatic Indicator 3: Health Sector Planning and Governance for Integrated People-Centred Services (HSG)-1.1: Availability of Clinical Guidelines

This indicator assesses whether up-to-date clinical guidelines for HTM and other primary health care services (e.g., ANC, immunization, child health) are available on-site. The rationale is that increased access to standardized clinical protocols funded through Global Fund grants is a prerequisite for delivering quality and consistent care, helping reduce variability in practices and improve clinical outcomes. Components involve physical verification of printed or digital copies of relevant national guidelines during the facility visit.

#### 8. Programmatic Indicator 4: RSSH/PP Medical Oxygen and Respiratory Care System (RCS)-1: Availability of Functional Oxygen Systems

This indicator determines whether oxygen delivery systems (e.g., cylinders, concentrators, pulse oximeters, flowmeters) are available and functional in a facility. The rationale recognizes the lack of standardized data across countries on oxygen functionality in routine information systems to monitor the COVID-19 related large-scale investments in oxygen as a lifesaving essential element in managing severe conditions (e.g., pneumonia as well as COVID-19), reflecting the facility’s capacity to respond to critical care needs.[14] Components include two key attributes: (i) availability of oxygen and delivery equipment (including functionality of flow meters for oxygen, humidifiers, and oxygen delivery apparatus); and (ii) availability of a functional pulse oximeter. Data are collected through direct observation.

#### 9. Programmatic Indicator 5: RSSH/PP Laboratory Systems (including national and peripheral): Availability of Essential Diagnostics

This indicator checks whether a facility/laboratory has a predefined set of essential diagnostic tests based on the WHO Essential Diagnostics List with a focus on HTM services. The rationale identifies diagnostic capacity as a critical bottleneck to accurate treatment and disease control. Components include a list of 27 essential diagnostic tests in facilities that have a laboratory and a list of 11 rapid and screening tests in facilities without a laboratory but that report conducting tests outside the facility. The list of tests is a sub-set of the WHO Essential Diagnostic List selected by the Global Fund lab specialists to inform their investments in HTM services. Data are collected through direct observation.

#### 10. Programmatic Indicator 6: RSSH/PP HRH-1: Vacancy Rate

This indicator measures the proportion of full-time funded but currently vacant HW positions at a facility. The rationale is that vacant posts signify systemic workforce shortages, which impair service delivery, reduce care quality, and limit a facility’s ability to implement integrated programming. Components include identifying approved positions versus those currently filled for physicians, nurses, midwives, lab technicians, pharmacists, and CHWs. Data are collected through interviews of the facility in-charge.

#### 11. Programmatic Indicator 7: RSSH/PP HRH-4: Provision of high quality HTM services by CHWs

This indicator measures the extent to which CHWs deliver high-quality HTM services in line with national protocols. The rationale reflects the critical role CHWs play in expanding access to essential services at the community level, while highlighting the need to assess not only their availability but also the quality of care they provide. Measurement is based on review of CHW service records and reporting forms to determine whether core tasks such as case assessment, diagnostic testing, treatment, referral, and health promotion are carried out. Data are collected through abstraction of CHW registers and monthly reports.

### Questionnaire Structure

The tHFA questionnaire has eight modules (Table 2), each designed for brevity and alignment with the 11 indicators. Screening questions on service availability (e.g., HIV treatment) in an early module allows for content filtering and quality-related questions for specific services (e.g., ART record review) in a subsequent module, which minimises respondent and data collector burden. The data collection methods chosen enable objectively verifiable data and do not require medically trained data collectors. All modules except patient exit interviews and CHW interviews incorporate the relevant and available questions from the HHFA. During the baseline survey, the questionnaire was pilot tested in field settings twice to refine the content and flow based on the understanding of the respondents and the interviewers.

**Table 2:** Summary of eight modules of the baseline Targeted Health Facility Assessment (tHFA) questionnaire.

| Module | Data collection method | Key components | Total number of questions | Number of questions from HHFA |
| --- | --- | --- | --- | --- |
| 1.Facility identifiers | Facility in-charge interview | Location, level of care | 16 | 14 |
| 2.Service availability & Readiness | Facility in-charge interview, direct observation | Services, staffing, guidelines, equipment | 81 | 22 |
| 3.Supportive supervision | Facility in-charge interview | Scope, frequency, content | 13 | 2 |
| 4.Patient-centeredness | Outpatient exit interview | Experience, respectful care, access challenges | 18 | 5 questions from WHO PREMs |
| 5.Support to CHWs | CHW interview | Support, supplies, remuneration | 30 | 0 |
| 5a. and 5b. CHWs Record review | CHW client register book | Quality of care | 31 | 0 |
| 6.Antenatal care | Record review | Service delivery record, tests, procedures | 20 | 17 |
| 7.HIV services | ART record review | Service delivery record, tests, procedures | 15 | 10 |
| 8.TB services | Record review | Service delivery record, tests, procedures | 8 | 8 |

### Sampling

A master facility list (MFL) of all health facilities in each country was obtained by the Global Fund and updated with information on ANC and HTM service availability. Any facility that offered only specialized services such as maternity homes or dental clinics was excluded from sampling. All public sector health facilities were included in the MFL of each country and private sector facilities were included in Madagascar, Gambia, Liberia, Madagascar, Guinea-Bissau, Mali, Cameroon, and Niger.

Each country was divided into three contiguous geographic domains, approximately equal in terms of population size and number of eligible primary health facilities. Facilities were then stratified by level of care provided: primary (health centres/posts), secondary (district/sub-district hospitals), and tertiary (referral/specialty hospitals) facilities. Altogether, 120 health facilities were selected using a two-stage sampling strategy (details below) in each country to balance representativeness at the national level with relevance for the Global Fund’s grant and program monitoring (Table 3)

**Table 3:** Sample size at country-level for the Targeted Health Facility Assessment (tHFA)

| Number of countries | Number of Domains | Number of facilities per domain (Stratified Random + Panel) | Total number of facilities in the baseline round |
| --- | --- | --- | --- |
| 18 | 3 | 28+12 | 84+36 |

Note: Exceptions to the 120 facilities per country sample size were: 1) Nigeria (N = 360 facilities, to allow for greater precision of sub-national indicator estimates) 2) Guinea-Bissau (N =114) and 3) Niger (N=118) as the rest of the eligible facilities in Guinea-Bissau and Niger were not accessible due to security reasons

In stage 1, in each geographic domain, starting with the tertiary, then secondary and primary facility strata, 30% of the sample (12 facilities per domain) was randomly sampled from facilities that had large patient volumes and received direct support from the Global Fund. These facilities constitute the panel for future rounds of the tHFA during the strategy period. In stage 2, from the remaining pool of eligible facilities in each domain (including those not sampled in stage 1), stratified random sampling was used to select the remaining 70% of the sample (28 facilities per domain). Sampled facilities that were inaccessible due to security reasons were replaced with a randomly sampled facility from the same domain at the same level of care (primary, secondary or tertiary). On average, less than 10% of the initially sampled facilities were replaced due to security reasons. Post-stratification weighting was used to account for the sampling method, including facility replacement and other sampling deviations (weights = 1/facility selection probability). This allows for the estimation of a 95% confidence interval of ± 10 percentage points for facility-level indicators and a difference of 16 percentage points (or higher) between 2 rounds of tHFA. In each health facility, random sampling was used to select five outpatients (for exit interview questions on patient experience of care), five associated CHWs (for interview questions on support to CHWs) and extract data from five records for ANC, ART, and TB clients (for questions on integrated quality of care). In summary, sampling in each country approximated a probability based selection of eligible health facilities and respondents.

### Ethical approvals

An institutional review board (IRB) in each country reviewed the tHFA questionnaire, the protocol, and consent forms and approved the data collection (Table A1 lists IRB authorities and reference numbers). In Madagascar, Nigeria, Gambia, and Uganda, the country-level IRB determined the assessment to be public health practice and therefore exempted it from human-subjects research. At every facility, the study obtained written consent from the facility in-charge, participating patients, and CHWs prior to interview or observation. In case of minor patients, the written consent was obtained from their parent/guardian. No personally identifiable information was collected from patients, CHWs, or clients included in the interviews or record reviews. All data were recorded in anonymized form, with no names or unique identifiers retained, and client sampling lists were destroyed immediately after record abstraction to ensure confidentiality.

### Data collection and quality assurance

The study collected all data in person, in-country, over four to six weeks, using tablet-assisted interviewing software (SurveyCTO; Dobility, Inc.; Cambridge, MA) to facilitate real-time quality checks and secure transmission. Two agencies managed the data collection: IQVIA (in 14 countries) and KPMG (in five countries) during the period of May 2024 to July 2025 (Table 4), to coincide with the signing of the Global Fund country grants, and the data collection for each country completed in 4-6 weeks.

**Table 4:** Data collection timelines in 18 countries (May 2024–July 2025)

| Sno | Country Name | Data Collection<br>(Start Date) | Data Collection<br>(End Date) |
| --- | --- | --- | --- |
| 1 | Burkina Faso | 31 May 2024 | 25 July 2024 |
| 2 | Cameroon | 17 July 2024 | 20 August 2024 |
| 3 | Chad | 18 August 2024 | 08 September 2024 |
| 4 | Congo | 15 July 2024 | 15 August 2024 |
| 5 | Congo (Democratic Republic) | 17 January 2025 | 07 March 2025 |
| 6 | Gambia | 05 June 2024 | 31 July 2024 |
| 7 | Ghana | 24 May 2024 | 11 July 2024 |
| 8 | Guinea-Bissau | 18 January 2025 | 28 February 2025 |
| 9 | Liberia | 16 May 2024 | 28 June 2024 |
| 10 | Madagascar | 21 May 2024 | 06 August 2024 |
| 11 | Malawi | 13 May 2024 | 12 June 2024 |
| 12 | Mali | 03 September 2024 | 05 October 2024 |
| 13 | Mozambique | 06 August 2024 | 04 September 2024 |
| 14 | Niger | 01 July 2024 | 09 November 2024 |
| 15 | Nigeria | 14 May 2024 | 23 July 2024 |
| 16 | Senegal | 16 April 2025 | 20 May 2025 |
| 17 | Tanzania (United Republic) | 23 September 2024 | 19 October 2024 |
| 18 | Uganda | 06 June 2024 | 19 July 2024 |

**Table 5:**
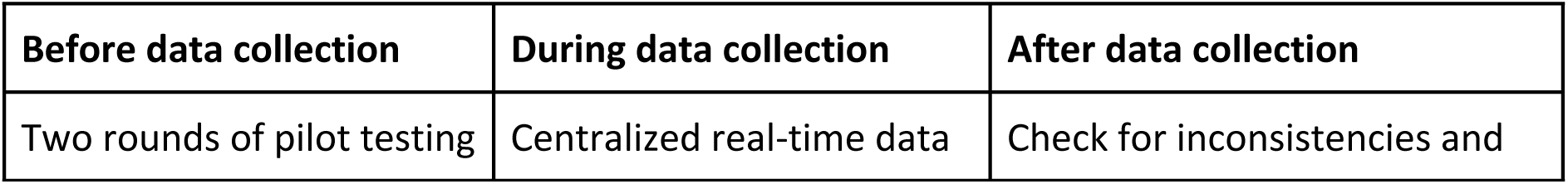
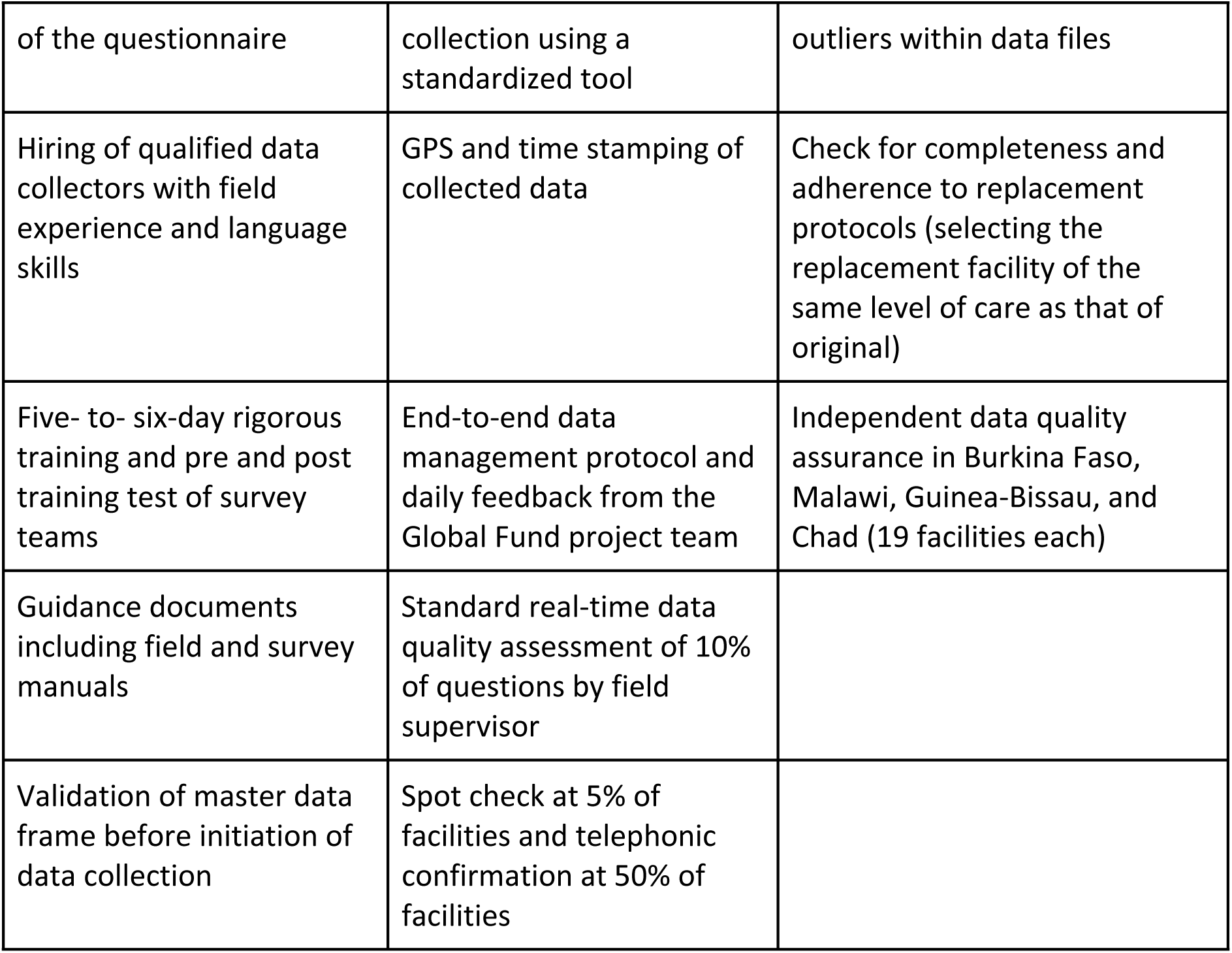
Data quality assurance for tHFA baseline implementation in 18 countries (May 2024–July 2025)

The average cost of data collection and database management for the tHFA in the 18 countries was US$ 70,000 per country (range of US $ 55,000 - 90,000).

Local data collectors with appropriate credentials, experience and language skills in each country underwent five to six days of training that included protocol review, questionnaire administration, field practice, research ethics (including written informed consent), privacy, and correct use of tablets. The team embedded comprehensive quality assurance (QA) throughout the tHFA implementation, through training, pilot-testing, and real-time monitoring, including before, during, and after data collection (Table 4). Independent data quality assurance (IDQA) was implemented in four of the 18 countries (Burkina Faso, Malawi, Guinea-Bissau and Chad) and data on a small sub-set of the tHFA questions were recollected in 19 of the 120 facilities within a week of the initial data collection. The agreement between question responses from the initial data collection and the IDQA was 80% or higher.

## Discussion

This paper describes the development and implementation of the baseline round of tHFA in 18 countries to monitor the Global Fund’s strategic objective of delivering IPCQS towards the goal of eliminating HTM. To our knowledge, tHFA is the first multi-country, nationally representative effort to monitor health service delivery processes and outcomes for the Global Fund grants and the national programs being implemented in these countries. By integrating insights on both disease-specific service delivery processes and system-level outcomes, the tHFA improves understanding of contribution of horizontal health systems strengthening investments to disease-specific programs. Institutionalized at the Global Fund, it responds to WHO’s IPCHS framework and the Lancet Global Health Commission’s call to increase the focus on service delivery processes and outcomes to improve the performance of health systems. It also addresses a gap in most health facility assessments by including CHW readiness and patient-reported experience, reinforcing the IPCHS principle of engaging communities and frontline providers.

Institutionally embedding the tHFA into the regular grant and strategic monitoring mechanisms of the Global Fund allowed for its sustainability and rapid implementation with timely data collection, specifically, by using existing processes for coordinating between technical and country teams and data collection agencies and ministries of health. The tHFA’s short, targeted set of indicators and reliance on methods of data collection (facility observation, interviews, and record review) that do not require surveyors to have specialized clinical knowledge helped optimize field costs. The use of a standardised, modular approach incorporating relevant questions from HHFA also allowed for comparability across settings while maintaining relevance to country-specific systems.

The targeted focus on measuring service delivery processes and outcomes, coupled with relatively low cost compared to existing HFAs, allowed the tHFA to be agile and enables repeated assessments over the five-year Global Fund strategy cycle for timely decision-making. The modular architecture provides the flexibility to add country-specific questions while maintaining KPI comparability. Close partnership with MoH through shared decision-making promotes mutual accountability and institutionalization, increasing the likelihood of sustained use for national planning, grant monitoring and incorporation of health systems outcomes into future grant applications. Future follow-up would determine the degree to which the methods and indicators are being adopted and financed by the country governments through inclusion into country grant proposals and other funding applications.

With recent reduction in the official development assistance (ODA) for Global Health, specifically US government funding for large scale health facility assessments (HFAs) like the SPA and HHFA, the tHFA can serve as cost-efficient platform for Global Health Institutions (GHI’s) to monitor investments through reliable data on service delivery processes and outcomes at the point of service delivery. The tHFA could facilitate alignment with the principles of mutual accountability, ownership and local leadership in health system strengthening efforts articulated in the Lusaka Agenda, a consensus document developed by stakeholders around the world regarding the future of the GHIs.[15] This collaborative approach aligns with IPCHS priorities on creating enabling environments and the Lancet Commission’s call for stronger health system leadership.[2,6]

The tHFA has limitations, however. First, the cross-sectional design does not allow measurement of changes in service integration and quality of care for a given patient over time, especially when management of conditions included in the assessment requires multiple visits to a health facility. The longitudinal measurement of the process and outcomes of care for pregnant women is part of the Maternal and Newborn Health (MNH) eCohort[16]and the Performance Monitoring and Accountability (PMA)[17] studies and could be included in the patient exit interviews for future rounds of tHFA. Second, the data on service integration and quality were abstracted from paper-based patient records, which may yield incomplete or incorrect information, compared to other methods of data collection, such as direct observation of service delivery, clinical vignettes, or mystery clients.[18,19]Available literature indicates an increase in availability of electronic health records in LMICs[20], use of which can improve the accuracy of estimates in future rounds. Moreover, where methodologically feasible, the paper-based data collection of tHFA can be complemented with faster methods for data collection, for example the Global Financing Facility (GFF) led Frequent Assessment & Health Systems tools for Resilience (FASTR) initiative[21] that collects data from the facility in-charge via phone interviews to identify and reduce service delivery constraints in PHC facilities. Finally, social desirability bias during the patient exit interview may cause over-reporting of service quality. Existing literature suggests that conducting phone-based exit surveys and collecting data after a few days of the facility visit can avoid this bias and could be implemented in future rounds.[22] Notwithstanding these limitations, given the dearth of data on IPCQS in these countries and a historical preponderance of input-based monitoring metrics in HFAs, the tHFA provides valuable data that can be used to improve performance of health systems in LMICs.

The progress on Global Fund’s strategic KPIs is based on change in tHFA indicators, therefore, we expect that any bias in measurement at baseline will remain constant when comparing 2 data sets collected the same way and in the same country at two points in time. Future rounds will require continued coordination between the Global Fund and the established national partners to maintain a core questionnaire and sampling consistency for KPI comparability, while retaining flexibility for country-specific priorities. In the face of constrained funding, the tHFA offers a scalable platform for GHIs to work with LMIC institutions in building a country-led, harmonized and efficient monitoring system for stronger health systems focused on mutual accountability to improve actual services delivery.

## Conclusion

The tHFA is a pragmatic, agile, scalable platform to monitor integrated service delivery and people-centred quality of care in LMICs. With a focused set of process- and outcome-oriented measures, inclusion of CHWs, and alignment with the Global Fund Strategy (2023–2028), it supports decision-making, programme monitoring, and health-system strengthening. By linking disease-specific delivery with system outcomes, it helps bridge the vertical–horizontal divide. Partnership with ministries of health fosters accountability, institutionalization and integration in national plans. Its modular design adapts to country priorities while maintaining KPI comparability, enabling repeated use for performance management and HSS tracking in a mutually accountable manner.

## Ethics Statement

Ethical approvals for the tHFA were obtained from institutional review boards (IRBs) in all participating countries. Details of national IRBs and approval reference numbers are provided in Table A1. In Madagascar, Nigeria, The Gambia, and Uganda, the assessment was determined to be public health practice and exempt from human-subjects research review. Written consent was obtained from all participants prior to interviews or observations.

## Data Availability Statement

The manuscript provides full methodological details of the Targeted Health Facility Assessment (tHFA). Copies of the tHFA questionnaire modules and related materials can be provided upon reasonable request to the corresponding author.

## Author Contributions

Conceptualization: S.G., S.M., M.M

Methodology: A.R., S.G., S.M., P.J, J.K.

Investigation: R.J.,J.K., P.J, A.A

Writing – original draft: S.G., J.K.

Writing – review & editing: S.M., C.W., R.R., P.J, A.A, M.M, A.R

Supervision: S.M., S.G

## Acknowledgments

We acknowledge the contributions of the Ministries of Health and data-collection partners across the 18 participating countries. We also thank colleagues from the Global Fund Secretariat for technical guidance and coordination support.

---

**Table A1:** IRB Approval authority and reference number by Country.

| Sno | Country Name | IRB Approval Authority | IRB Reference number |
| --- | --- | --- | --- |
| 1 | Burkina Faso | Health Research Ethics Committee | DELIBERATION N° 2024-04-89 |
| 2 | Cameroon | National Ethical Committee For Research On Human Health | N° 2024/04/4660/CE/CNERSH/SP |
| 3 | Chad | National Bioethics Committee of Chad (CNBT) | N036/MESRSFP/SE/SG/CNBT/SG/2024 |
| 4 | Congo | Institutional Ethics Committee – FCRM | Avis n° 054/CEI/FCRM/2024 |
| 5 | Congo (Democratic Republic) | University of Kinshasa School of Public Health Ethics Committee | ESP/CE/209/2024 |
| 6 | Gambia | Ethical Approval - Not Required |  |
| 7 | Ghana | Ghana Health Service Ethics Review Committee | GHS-ERC: 024/03/24 |
| 8 | Guinea-Bissau | National Committee on Health Research Ethics (CNES) | N° Ref. 159/CNES/INASA/2024 |
| 9 | Liberia | Atlantic Center for Research and Evaluation Institutional Review Board (ACRE IRB) | FWA00032198 |
| 10 | Madagascar | Ethical Approval - Not Required |  |
| 11 | Malawi | National Health Sciences Research Committee ((NHSRC) | MED/4/36c approval number 4324 |
| 12 | Mali | Ethics Committee for Health and Life Sciences (CNESS) | Décision n° 2024 006 /MSDS-CNESS du 26 JUIN 2024 |
| 13 | Mozambique | National Bioethics Committee for Health | 427/CNBS/24 |
| 14 | Niger | National Ethics Committee for Health Research (CNERS) | DELIBERATION N°29/2024/CNERS |
| 15 | Nigeria | Ethical Approval - Not Required |  |
| 16 | Senegal | National Ethics Committee for Health Research (CNERS) | 00000070/MSAS/CNERS/SP |
| 17 | Tanzania (United Republic) | National Institute for Medical Research | NIMR/HQ/R.8a/Vol.IX/4683 |
| 18 | Uganda | Ethical Approval - Not Required |  |

## References

Bitton A, Ratcliffe HL, Veillard JH, Kress DH, Barkley S, Kimball M, et al. Primary health care as a foundation for strengthening health systems in low-and middle-income countries. J Gen Intern Med. 2017;32: 566–571. doi:10.1007/s11606-016-3898-5

Kruk ME, Gage AD, Arsenault C, Jordan K, Leslie HH, Roder-DeWan S, et al. High-quality health systems in the Sustainable Development Goals era: time for a revolution. The Lancet Global Health. Elsevier Ltd; 2018. pp. e1196–e1252. doi:10.1016/S2214-109X(18)30386-3

World Health Organization, World Bank. Tracking universal health coverage: first global monitoring report. Geneva; 2015. Available: https://www.who.int/publications/i/item/9789241564977

Leslie HH, Ndiaye Y, Kruk ME, others. Effective coverage of primary care services in eight high-mortality countries. BMJ Glob Health. 2017;2: e000424.

World Health Organization. World health report 2013: Research for universal health coverage. 2013. Available: https://iris.who.int/handle/10665/85761

World Health Organization. Framework on integrated, people-centred health services: report by the Secretariat. Sixty-ninth World Health Assembly, A69/39, provisional agenda item 161. Geneva: World Health Organization; 2016. Available: https://apps.who.int/gb/ebwha/pdf_files/WHA69/A69_39-en.pdf?ua=1&ua=1

World Health Organization, United Nations Children’s Fund. Operational Framework for Primary Health Care: transforming vision into action. Geneva: World Health Organization; 2020. Available: https://iris.who.int/bitstream/handle/10665/337641/9789240017832-eng.pdf

World Health Organization. Declaration of Astana. Geneva; 2019. Available: https://iris.who.int/bitstream/handle/10665/328123/WHO-HIS-SDS-2018.61-eng.pdf

The Global Fund. Fighting Pandemics and Building a Healthier and More Equitable World: Global Fund Strategy (2023–2028). Geneva: The Global Fund to Fight AIDS, Tuberculosis and Malaria; 2021. Available: https://www.theglobalfund.org/media/11612/strategy_globalfund2023-2028_narrative_en.pdf

The Global Fund. Key Performance Indicators (KPIs) Handbook for the 2023-2028 Strategy. 2023. Available: https://www.theglobalfund.org/media/12681/strategy_globalfund2023-2028-kpi_handbook_en.pdf

Asefa A, Dossou JP, Hanson C, Hounsou CB, Namazzi G, Meja S, et al. Methodological reflections on health system-oriented assessment of maternity care in 16 hospitals in sub-Saharan Africa: an embedded case study. Health Policy Plan. 2022;37: 1257–1266. doi:10.1093/heapol/czac078

The Global Fund. Modular Framework Handbook. 2023 May. Available: https://resources.theglobalfund.org/media/13907/cr_modular-framework_handbook_en.pdf

Rowe AK, Rowe SY, Peters DH, Holloway KA, Chalker J, Ross-Degnan D. Effectiveness of strategies to improve health-care provider practices in low-income and middle-income countries: a systematic review. Lancet Glob Health. 2018;6: e1163–e1175. doi:10.1016/S2214-109X(18)30398-X

Ijaz N, Lee T, Furtado N, Macher E, Muitheri Z, Park BJ, et al. A rapid facility-level assessment of oxygen systems in 39 low-income and middle-income countries: a cross-sectional study. Lancet Glob Health. 2025;13: e646–e655.

Future of Global Health Initiatives. The Lusaka Agenda: Conclusions of the Future of Global Health Initiatives Process. Future of Global Health Initiatives; 2023. Available: https://d2nhv1us8wflpq.cloudfront.net/prod/uploads/2023/12/Lusaka-Agenda.pdf

Arsenault C, Wright K, Taddele T, Tadele A, Derseh Mebratie A, Tiruneh Tiyare F, et al. The maternal and newborn health eCohort to track longitudinal care quality: study protocol and survey development. Glob Health Action. 2024;17: 2392352.

Performance Monitoring for Action (PMA). PMA Data. In: Baltimore (MD): Johns Hopkins Bloomberg School of Public Health [Internet]. [cited 25 Sep 2025]. Available: https://www.pmadata.org/

Martin SK, Hall MAK, Molitch-Hou E, Benaderet A, Park JJ, Sweet M. Case to Vignette: A Framework to Elevate Narrative by Embracing Vignette-Based Learning in Medicine. J Gen Intern Med. 2025 [cited 7 Sep 2025]. doi:10.1007/s11606-025-09531-5

Fitzpatrick A, Tumlinson K. Strategies for optimal implementation of simulated clients for measuring quality of care in low-and middle-income countries. Glob Health Sci Pract. 2017;5: 108–114.

Ngugi P, Babic A, Kariuki J, Santas X, Naanyu V, Were MC. Development of standard indicators to assess use of electronic health record systems implemented in low-and medium-income countries. PLoS One. 2021;16: 1–15. doi:10.1371/journal.pone.0244917

The Global Financing Facility and the World Bank Group. FASTR : The GFF’s approach to rapid cycle analytics and data use. 2024 [cited 3 Oct 2025]. Available: https://data.gffportal.org/key-theme/FASTR

Shamebo D, Derseh Mebratie A, Arsenault C. Using an Interactive Voice Response Survey to Assess Patient Satisfaction in Ethiopia: Development and Feasibility Study. JMIR Form Res. 2025;9: e67452. doi:10.2196/67452

